# Intersectional consequences for marginal fairness in prediction models for emergency admissions

**DOI:** 10.1101/2024.11.05.24316769

**Authors:** Elle Lett, Shakiba Shahbandegan, Yuval Barak-Corren, Andy Fine, William G. La Cava

## Abstract

**Background:** Fair clinical prediction models are crucial for achieving equitable health outcomes. Recently, intersectionality has been applied to develop fairness algorithms that address discrimination among intersections of protected attributes (e.g., Black women rather than Black persons or women separately). Still, the majority of medical AI literature applies marginal de-biasing approaches, which constrain performance across one or many isolated patient attributes. We investigate the extent to which this modeling decision affects model equity and performance in a well-defined use case in emergency medicine.

**Methods:** The study focused on predicting emergency room admissions using electronic health record data from two large U.S. hospitals, Beth Israel Deaconess Medical Center (MIMIC-IV-ED, n=160,016) and Boston Children’s Hospital (BCH, n=22,222), covering both adult and pediatric populations. In a comprehensive experiment over fairness definitions, modeling methods, we compared the performance of single- and multi-attribute, marginal de-biasing approaches to intersectional de-biasing approaches.

**Results:** Intersectional de-biasing produces greater reductions in subgroup calibration error (MIMIC-IV: 21.2%; BCH: 27.2%) than marginal de-biasing (MIMIC-IV: 10.6%; BCH: 22.7%), and also lowers subgroup false negative rates on MIMIC-IV an additional 3.5% relative to marginal de-biasing. These fairness gains were achieved without a significant decrease in model accuracy between baseline and intersectionally-debiased models (MIMIC-IV: AUROC=0.85*±*0.00, both models; BCH: AUROC=0.88*±*0.01 vs 0.87*±*0.01). Intersectional de-biasing more effectively lowered subgroup calibration error and FNRs in low-prevalence groups in both datasets compared to other de-biasing conditions.

**Conclusion:** Intersectional de-biasing better mitigates performance disparities across intersecting groups compared to marginal approaches for emergency admission prediction. These strategies meaningfully reduce group-specific error rates without compromising overall accuracy. These findings highlight the importance of considering interacting aspects of patient identity in model development, and suggest that intersectional de-biasing would be a promising gold standard for ensuring equity in clinical prediction models.

## INTRODUCTION

Emergency departments (EDs) are dynamic environments where patients present with varying acuity, requiring tailored and efficient treatment plans that prioritize achieving desired health outcomes while optimizing clinician workflow and hospital resource utilization. EDs often function as safety-net care for marginalized populations with reduced economic resources or healthcare access, particularly among minoritized ethnoracial groups in the United States^1^. These populations also experience the most severe health inequities in disease burden, mortality, and morbidity^2–4^. Racialized health inequities also manifest throughout the ED workflow; Black and Hispanic/Latino patients are subject to longer wait times for initial evaluation by a physician in the ED^5^, despite data suggesting that Black and Hispanic individuals account for a disproportionate amount of ED visits and are more likely to be repeat visitors^1^. After initial triage, Black patients who are designated for admission also experience longer ED boarding times (stays in the ED before entering an inpatient service)^6^, with such delays associated with adverse health outcomes including intensive care unit (ICU) mortality rates^7^ and ventilator-associated pneumonia^8^.

### Machine Learning Models Capacity for Improving Emergency Department Patient Management

A key challenge in ED workflow contributing to wait time inequities is coordinating admissions for patients needing inpatient care, as hospital beds are a limited resource. Bed coordination - the assignment of patients to care teams and beds - can create bottlenecks, increasing ED boarding times and delaying treatment when demand exceeds capacity or allocation is inefficient. Machine learning (ML) models can help accelerate this process by identifying potential admissions early during triage and initial work-up, before the formal decision to admit is made (Fig. 1). Our study builds on previous ED admission prediction models that have shown strong performance in adult^9^ and pediatric^10^ settings, improving and complementing the assessments of patient disposition made by attending physicians^11^.

**Figure 1:**
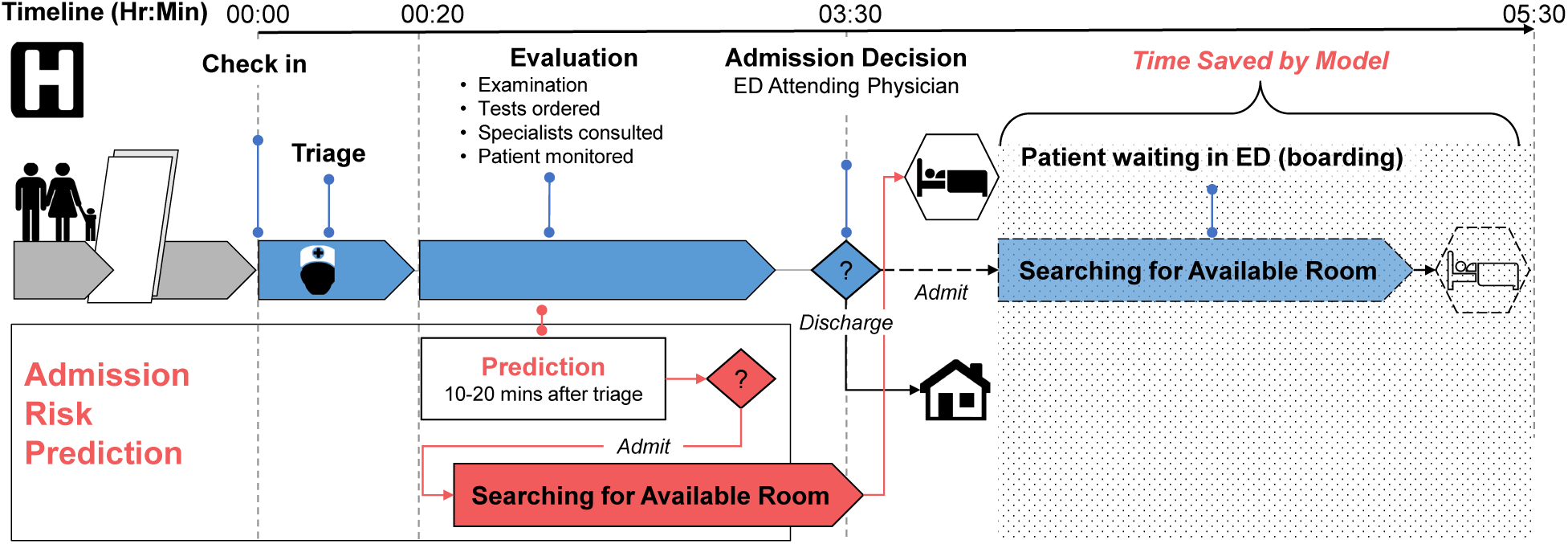
An illustration of the admission prediction task and its utility in the emergency department (ED) during the typical timeline of a patient visit. Normally, patients who will be admitted wait while care coordinators find an available room (known as boarding). Admission prediction algorithms flag high risk patients early in the visit so that the bed coordination can happen before the ED attending physician makes an admission decision for the patient.

### Fairness and Intersectionality

Previous ED admission prediction models focused on optimizing overall performance without addressing differences in subgroup outcomes. Given existing inequities in ED wait and board times, a “fairness-agnostic” model could narrow, maintain, or even widen disparities between privileged and marginalized groups. Therefore, we develop “fairness-aware” models that optimize both overall accuracy and equitable performance across groups defined by demographic traits. Prior work in fair ML has described the common limitation of many fairness approaches to focusing on groups defined by a single demographic trait such as race, or considering multiple demographic traits in isolation (i.e. race and gender separately)^12^. We refer to these approaches as “marginal”, as they focus on the marginal distribution of one or more protected attributes while ignoring groups defined by their intersections. Marginal fairness approaches are subject to “fairness gerrymandering“^13,14^, wherein models that are “fair” for groups defined by single attributes (i.e. Black people, or women, separately) still exhibit unfair performance for groups defined by intersections of protected attributes (i.e. Native American women, or Latino men). We provide a break-down of currently available fair ML algorithms and their support for intersecting subgroup definitions in Table 1.

**Table 1:** Properties of a number of algorithms proposed for fair machine learning, along with their properties and support for intersectional fairness definitions. DP: Demographic Parity; FNR: False Negative Rate; FPR: false positive rate. Model-Agnostic indicates that the algorithm supports many common base ML models. The algorithms in bold are the two used in this study.

| Stage | Algorithm | Fairness Definition |  |  | Intersectional Groups | Model-Agnostic |
| --- | --- | --- | --- | --- | --- | --- |
|  |  | DP | FNR/FPR | Calibration |  |  |
| Pre | Reweighting<sup>15</sup> | ✓ |  |  |  | ✓ |
|  | Fair Feature Selection<sup>16</sup> | ✓ |  |  |  | ✓ |
| Train | Adversarial Debiasing<sup>17</sup> |  | ✓ |  |  |  |
|  | Differential Fairness<sup>18</sup> | ✓ |  |  | ✓ |  |
|  | Exponentiated Gradients<sup>19</sup> | ✓ | ✓ |  |  | ✓ |
|  | GerryFair<sup>13</sup> | ✓ | ✓ |  | ✓ | ✓ |
|  | **FOMO**<sup>20</sup> | ✓ | ✓ | ✓ | ✓ | ✓ |
| Post | Threshold Optimization<sup>21</sup> | ✓ | ✓ |  | ✓ |  |
|  | Calibrated Equalized Odds<sup>22</sup> |  | ✓ | ✓ |  | ✓ |
|  | **MultiCalibration**<sup>23</sup> |  |  | ✓ | ✓ | ✓ |
|  | MultiAccuracy<sup>24</sup> |  | ✓ |  | ✓ | ✓ |

Approaches to mitigate fairness gerrymandering are rooted in intersectionality, a framework established by legal scholar Kimberlé Crenshaw^25,26^ and sociologist Patricia Hill Collins^27^, but with origins in 1830s social movements^28–30^. Intersectionality views systems of oppression such as racism and cis-sexism as co-occurring, emphasizing that analyzing a single axis of discrimination—such as race—fails to capture the harms experienced by individuals facing multiple forms of discrimination^31^. Our previous work shows how this framework applies to ML fairness throughout different stages of the prediction task, from defining to evaluation and updating the task^12^.

In algorithmic fairness, this framework motivates what we refer to as “intersectional” fairness that constrains model performance across groups that are defined by the intersections of protected attributes, rather than what we refer to as “marginal” fairness that is only concerned with the groups defined by the marginal distributions of one or more protected attributes. Theoretically, intersectional fairness is clearly ideal; in practice it can be difficult to achieve computationally due to scarce data on multiply-marginalized groups.

### De-biasing and Evaluating Fairness

Fairness metrics must be selected based on specific context of the implementation environment and adapted to the prediction task^12^. Depending on the hospital’s patient population, ED traffic, and operating practices, different metrics may be most salient to optimizing care across groups. For example, in an ED with particularly high ethnoracial inequities in boarding wait times, ensuring fair calibration would ensure that specific groups aren’t systematically deprioritized or over-prioritized by the algorithm via assigned risk scores. Ensuring low subgroup false negative rates (FNRs), meanwhile, would help ensure that no one group is being falsely discharged at a higher rate. To cover the breadth of potential use case scenarios we focus on two fundamental notions of fairness: *sufficiency*, i.e. patients with the same risk score should experience outcomes at a rate that is independent of group membership; and *separation*, i.e. patients with the same outcomes should receive risk scores that are independent of group membership^32^. For example, if an ED admission model meets sufficiency, patients with a 90% risk score should have equal admission likelihoods regardless of group membership. Conversely, if the model meets separation, risk scores for admitted patients should not differ by group, meaning false negative rates (FNRs) and false positive rates (FPRs) should be the same across groups. Both of these traits, sufficiency and separation, are important characteristics for fair prediction models, yet cannot be simultaneously satisfied when admission rates differ among groups^33^. Hence, we study both notions here by applying two fairness algorithms: one that achieves sufficiency by de-biasing group-level calibration, and one that achieves (a relaxation of) separation by de-biasing group-level FNRs. The first algorithm, multicalibration boosting^34^, is a post-processing algorithm that constrains the group-level calibration error. The second, fairness-oriented multiobjective optimization (FOMO)^20^, is a training algorithm we use to constrain group-level FNRs.

In our experiments, we evaluate the ED admission prediction task (Fig. 1) across adult and pediatric populations in two Boston-based healthcare centers.With these models, we compare the performance of marginal and intersectional de-biasing approaches with multicalibration and FOMO, specifically with 1) no de-biasing, 2) marginal de-biasing based on single-attributes (ethnoracial group or gender) or multiple attributes concomitantly (ethnoracial group and gender), and 3) intersectional de-biasing based on ethnoracial group and gender. We implement these de-biasing approaches on both logistic regression and random forest base models. The overall goal of the present study is to measure the extent to which optimization of algorithmic fairness on marginal groups transfers to intersectional patient groups, under different definitions of fairness, models, and clinical settings.

## METHODS

### Data Curation

We base our experiments on the task of inpatient hospital admission prediction for patients visiting the ED. Recently, multiple care centers have sought to develop, validate, and deploy ML models for this task, due to its significant impact on patient flow.^9–11,35^ In our experiments we use data from two EDs that are described in detail in Table 2. The first is from the Medical Information Mart for Intensive Care-IV Emergency Department (MIMIC-IV-ED) database^36^, a freely available data source on ED visits to Beth Israel Deaconess Medical Center between 2011 and 2019. The second is collected from Boston Children’s Hospital (BCH) ED from 2017 to 2018. After data preprocessing (see Supplement for more details), our analysis consists of 160,016 visits by 90,005 unique patients in the MIMIC-IV cohort and 22,222 visits by 17,938 unique patients in the BCH cohort.

**Table 2:**
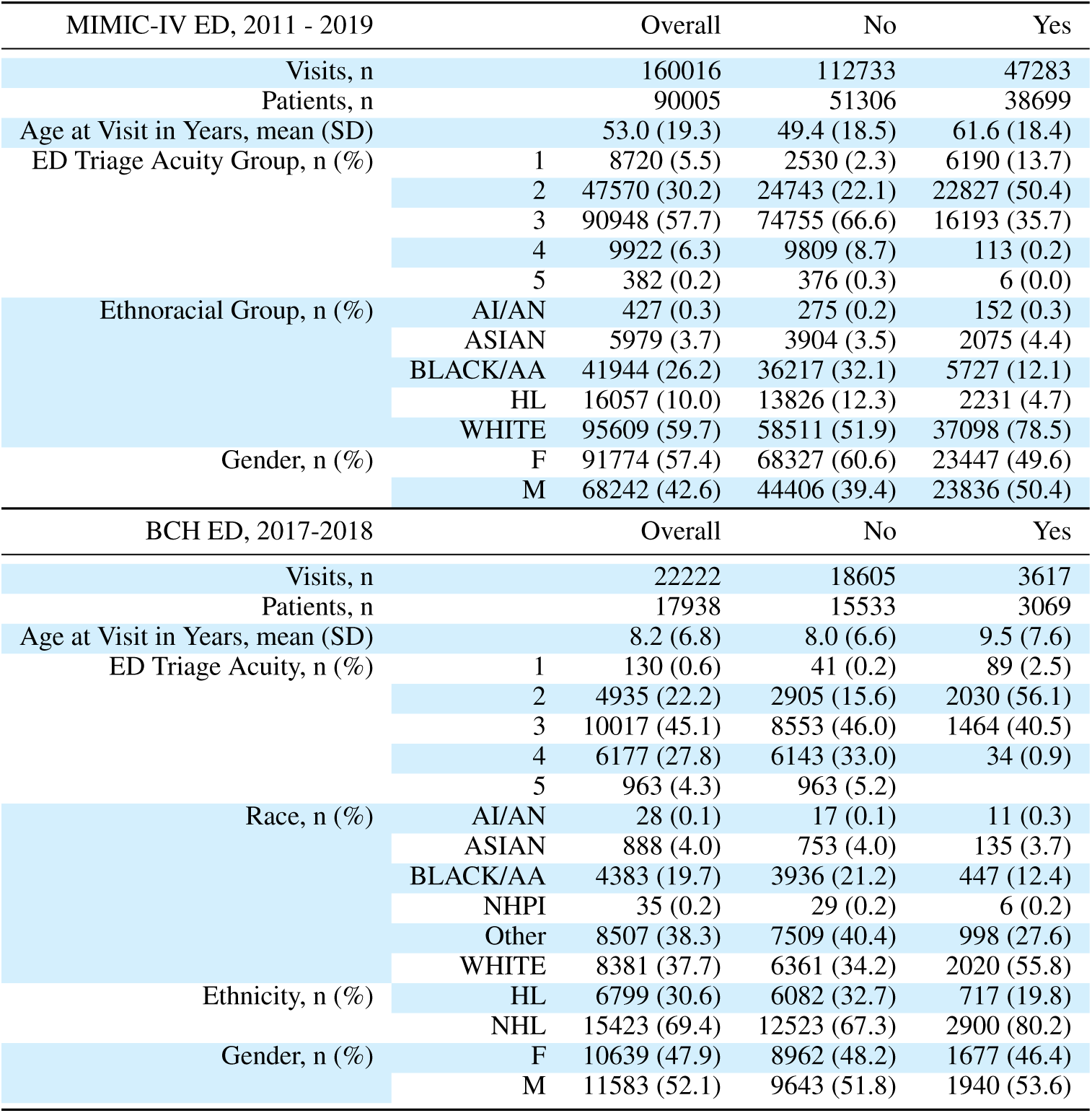
Patient visit characteristics for the MIMIC-IV and BCH data. AI/AN: American Indian / Alaskan Native; AA: African American; NHPI: Native Hawaian Pacific Islander; (N)HL: (Not) Hispanic/Latino; F: Female; M: Male.

### Model Development

In both cohorts, we train a model to predict admission to an in-patient service among patients whose final disposition has yet to be decided. We use data collected during check-in (e.g. chief complaint), triage (e.g. vitals), patient clinical history (e.g. number of previous admissions) and demographic data. In the BCH cohort, we include additionally available data collected during the first 60 minutes of a patient’s stay, including lab orders and medications. Table 3 lists the full set of features used in both cohorts.

**Table 3:** Features used for Emergency admission prediction in the MIMIC-IV and BCH cohorts. The BCH data includes a larger set of predictors (n = 155, BCH; n = 60, MIMIC-IV) including indicators of laboratory tests and a larger set of reported symptoms beyond chief complaint. HR: heart rate; RR: respiratory rate; SBP: systolic blood pressure; DBP: diastolic blood pressure; BMI: body mass index.

| Description | Features |
| --- | --- |
| MIMIC-IV |  |
| Vitals | temperature, HR, RR, oxygen saturation, SBP, DBP |
| Triage Acuity | Emergency Severity Index<sup>37</sup> |
| Check-in Data | chief complaint, self-reported pain score |
| Health Record Data | no. previous visits, no. previous admissions |
| Demographic Data | ethnoracial group, gender, age, marital status, insurance, primary language |
| BCH |  |
| Demographic Data | Gender, Race, Ethnicity, Age |
| Check-in Data | ED Day Of Week, ED Checkin Month, ED Checkin Year, ED Arrival Mode, Weekend, Miles Traveled, Patient State of Residence |
| Triage Data | Emergency Severity Index, ED Teams, ED Room Type, chief complaint, pain count, pain max, pain increase, CHEWS<sup>38</sup> count, CHEWS max, CHEWS increase |
| Medications | acetaminophen, albuterol, dexamethasone, epinephrine/lidocaine/tetracaine topical, ibuprofen, ipratropium, ondansetron, Sodium Chloride 0.9%, gastro count, gastro max, gastro increase, time till first med, drugs count, medication route (inhaled, intravenous, nebulized, oral, topical) |
| Vitals | HR count, HR min, HR max, HR sd, HR mean, RR count, RR min, RR max, RR sd, RR mean, temp low, temp normal, temp high low, temp not taken |
| Lab Test Orders | labs count, time till first lab, Blood Culture Routine, Aerobic, Blood Culture, Aerobic and Anaerobic, Blood Gas, Venous, C-Reactive Protein, Calcium, Plasma, Chemistry Panel, Chemistry Extended Panel, Complete Blood Count with Differential, Differential, Automated, Drug Screen, Urine (drugs of abuse), Erythrocyte Sedimentation Rate, Lipase, Plasma, Liver Function Tests, Urinalysis, Dipstick, Urine Culture, time till first test, tests count, Chest X-ray |
| Symptoms | Abdominal pain, Attention deficit disorder with hyperactivity, Allergic rhinitis, Anxiety, Asthma, Atopic eczema, Autistic disorder, Chronic constipation, Constipation, Cough, Dental caries, Depression, Dermatitis, Developmental delay, Dysphagia, Epilepsy, Eustachian tube dysfunction, Feeding problem, Fever, Food allergy, Gastroesophageal reflux, Global developmental delay, Headaches, Hypotonia, Obesity, Obstructive sleep apnea, Oropharyngeal dysphagia, Other Problem, Patent ductus arteriosus, Patent foramen ovale, Pediatric BMI greater than or equal to 95th percentile, Prematurity, Recurrent acute otitis media, Seizure, Snoring, Speech delay, Tonsillar and adenoid hypertrophy, Tonsils hypertrophy, Vitamin D deficiency, Vomiting |
| Clinical History | problems count, prior visits, prior admissions, admission ratio |

We test two baseline ML models: tree ensembles implemented in XGBoost (main tables and figures) and penalized logistic regression models (see Supplement). The hyperparameters of these models were tuned via halving grid search.

### Fairness Approaches

For all models, we experiment with multicalibration post-processing to improve subgroup calibration performance and fairness-oriented multiobjective optimization (FOMO) to improve subgroup FNRs.

#### Multicalibration Postprocessing

Multicalibration post-processing^23,34^ allows for flexible specification of groups for marginal and intersectional fairness models. Briefly, assume we have sample data (**x***_i_, y_i_*), where **x***_i_* is a vector of features and *y* is a binary outcome for individual *i*, drawn from joint distribution *D*. Let *C* represent a collection of subsets specified by protected attributes in **x** (i.e., subgroups). An *α*-multicalibrated model fulfills the constraint that among all subsets in *C* and binned prediction intervals, the absolute difference between the expected outcome and expected prediction is at most *α*. Hébert-Johnson^23^ showed that multicalibration is achieved without a fairness-utility tradeoff such that multicalibrated models have at least the same predictive power as the base model, which is ideal for our prediction task. The multicalibration algorithm updates model predictions until all groups defined by binned prediction intervals within collections in *C* with group probability greater than *γ* satisfy the calibration error constraint *α*. For our main results we used *α*=0.01 (constrain calibration error to 0.01) and *γ*=0.001 (consider groups with 0.1% or higher probability). The supplement contains a sensitivity analysis of these hyperparameters.

#### Fairness-Oriented Multiobjective Optimization

Achieving different notions of fairness in machine learning involves balancing the tradeoff between error and fairness, where increased fairness may lead to higher error rates, and vice versa. Traditionally, fair machine learning methods treat this as a single objective problem, introducing a parameter to weigh error against fairness. FOMO optimizes this tradeoff through multi-objective optimization, treating error and fairness as separate objectives^20^.

We use FOMO to jointly optimize the overall balanced accuracy of the models while minimizing the maximum FNRs among intersectional subgroups. This fairness definition has two motivations: first, it assumes that false discharges from the emergency room have the potential to cause more harm to a patient than a false admission. Second, unlike fairness metrics that optimize for FNR parity among groups, which can be achieved e.g. by making the model worse for some subgroups where it performs well, this metric focuses solely on improving the worst-case performance among patient subgroups. Minimizing subgroup FNRs must be balanced with minimizing overall FNRs and overall FPRs, which cause distributed harm to waiting patients due to overcrowding; hence, we jointly maximize for overall balanced accuracy.

#### Protected Attributes and Intersectionality

The experiment in this study focuses on three protected attributes: race, ethnicity, and gender (in the MIMIC-IV cohort, race and ethnicity are reported as a combined ethnoracial variable). We observe stark differences in admission rates by intersections of race, ethnicity, and gender (See Table S1), suggesting the importance of a performance-based fairness constraint (e.g., calibration or error rates) as opposed to demographic parity, which would cause substantial deviations in subgroup admission rates.

#### Statistical Tests

All reported *p*-values are the result of two-sided Mann-Whitney-Wilcoxon tests with Holm-Bonferroni correction.

## RESULTS

### Fairness without accuracy tradeoffs

The prevailing understanding of fairness as derived from the notions of equalized odds and demographic parity is that they require trade-offs with overall accuracy^22^.This trade-off is theoretically well-established, yet recent work has shown that in practice, such trade-offs may be negligible^39^. Our findings were consistent with the latter: across both data sets (MIMIC-IV and BCH), and both fairness targets (calibration and FNR), de-biasing on gender, ethnoracial identity, both concomitantly (marginally), and across intersectional groups, had nearly identical overall classification performance (mean AUROC within *±*0.01; Fig. 2). When tasked with balancing FNRs on the BCH cohort, intersectionally de-biased models exhibited slightly lower area under the precision recall curve (base scenario AUPRC: 0.67*±*0.01; intersectional scenario AUPRC: 0.64*±*0.02) due to lower precision in operating regimes with low recall/sensitivy, but nearly identical precision for model operating points with moderate to high sensitivity/recall that are desirable in this use case (Fig. 2, bottom right curve).

**Figure 2:**
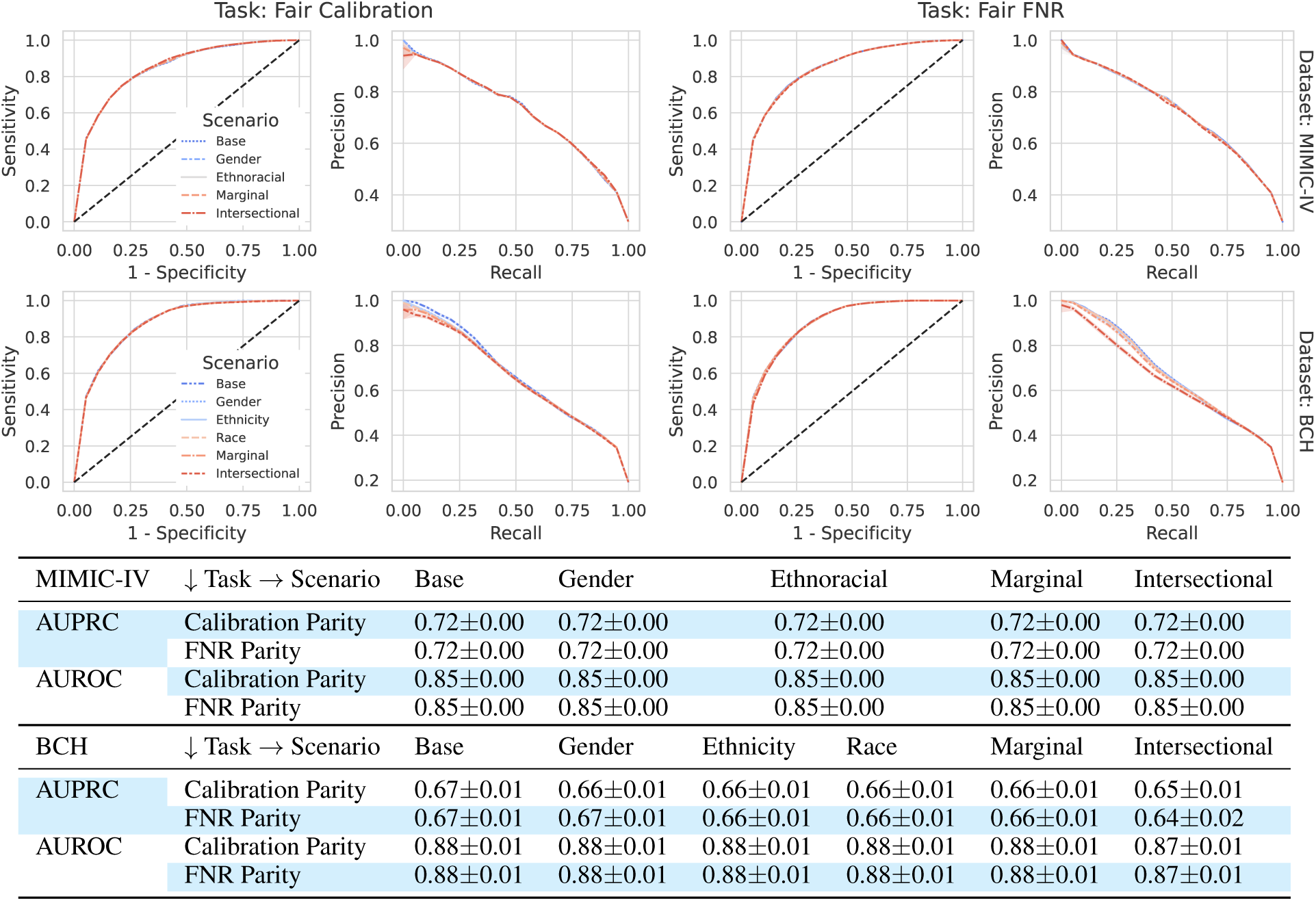
De-biased models perform as well as baseline models. **(a)** Receiver operating characteristic (ROC) curves and precision-recall curves for the prediction models on data from MIMIC-IV (top row) and BCH (bottom row). The left and right columns of subplots compare debiasing scenarios for fair calibration and fair false negative rates (FNR), respectively. **(b)** The mean (*±* standard deviation) area under the ROC curve (AUROC) and precision-recall curve (AUPRC) of prediction models by dataset, fairness task, and modeling scenario, corresponding to the curves above. In general, the fairness-aware models perform very similarly to the baseline models.

### Fairness gains with intersectional de-biasing

To compare fairness-unaware, marginal single-attribute, marginal multi-attribute, and intersectional de-biasing approaches at a high level, we compare the expected calibration error (ECE) and FNRs for the intersectional groups (ethnoracial group and gender cross-strata) across de-biasing conditions in Fig. 3. We observe that multi-attribute, marginal fairness de-biasing reduces ECE among intersectional groups on MIMIC-IV and BCH by 10.6% and 22.7%, whereas the fully intersectional approach reduces ECE by 21.2% and 27.2%, respectively (Fig. 3 left). In a similar vein, intersectional fairness de-biasing results in significantly lower FNRs among intersectional groups in the cohort compared to baseline (11% reduction, MIMIC-IV, *p* < 1e-16; 6.4% reduction, BCH, *p* < 3e-6). On MIMIC-IV, intersectional de-biasing reduces intersectional FNRs by an additional 3.5% compared to marginal fairness de-biasing (*p* = 1e-5). We observe across the experimental results that de-biasing on ethnoracial group produces a larger singular reduction in error rates among intersectional groups than de-biasing on gender alone, but that de-biasing using the intersectional combination of ethnoracial group and gender yields better performance than considering either attribute alone, or additively.

**Figure 3:**
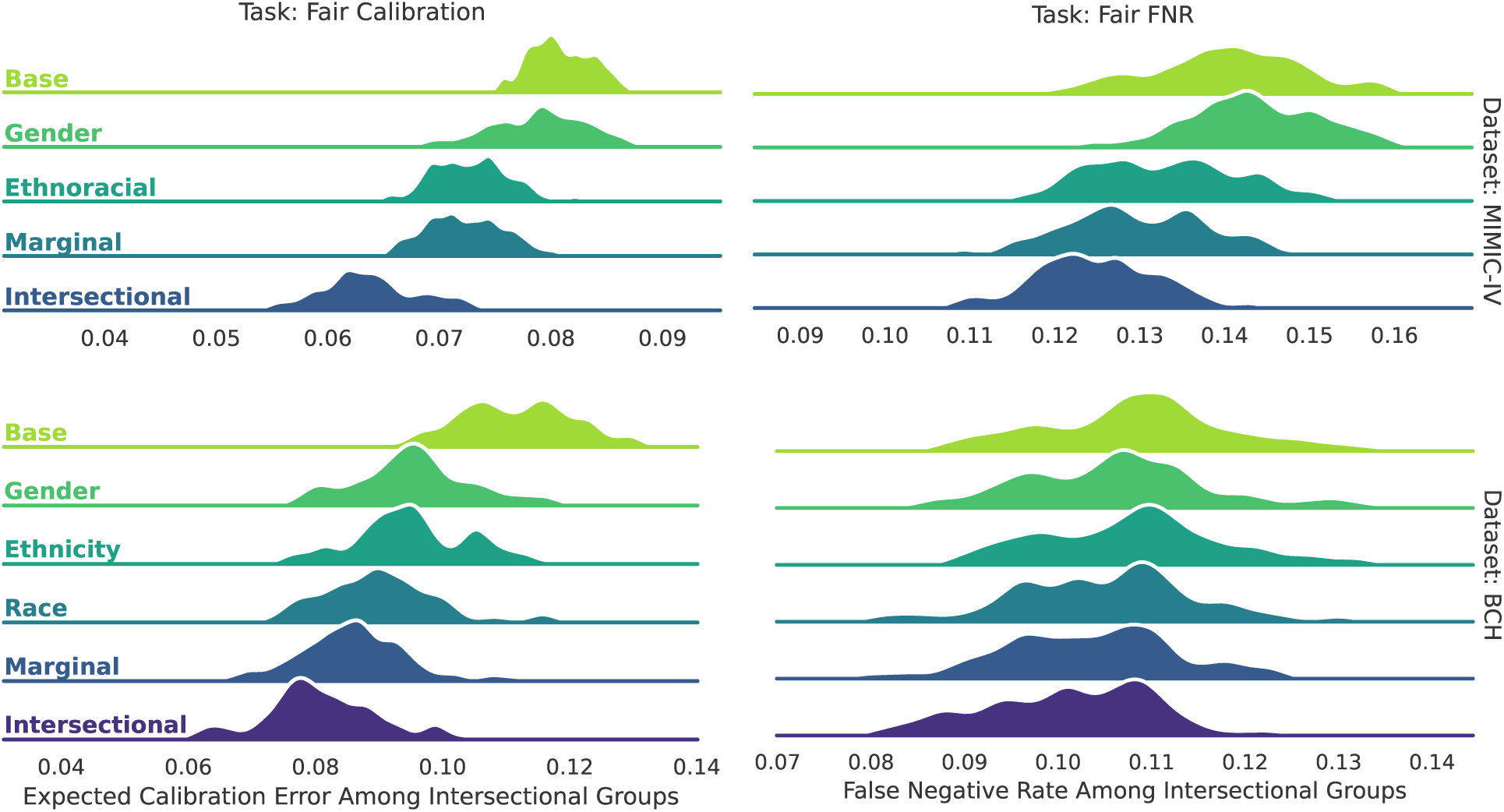
Intersectionally de-biased models improve fairness for intersectional groups beyond marginally de- biased models. Fairness measures under different de-biasing scenarios for MIMIC-IV (top) and BCH (bottom). Left plots report the expected calibration error (ECE) among intersectional groups when trying to ensure within-group calibration. Right plots report false negative rate among intersectional groups when optimizing for equal group-wise false negative rates. The scenarios (Base, Intersectional, etc.) are detailed in Table 4.

**Table 4:** Experimental setup for assessing algorithmic fairness under intersectional and marginal fairness scenarios.

| Variable | Settings |
| --- | --- |
| Group Construction Scenarios | **Base:** no fairness optimization<br>**Gender, Race, Ethnicity, Ethnoracial:** protect single attribute<br>**Marginal:** marginally protect all sensitive attributes<br>**Intersectional:** protect intersections of all sensitive attributes |
| Fairness Task (Algorithm) | Fair Calibration (Multicalibration Boosting)<br>Fair False Negative Rate (Fairness-Oriented Multiobjective Optimization) |
| Base Classifier | Penalized Logistic Regression (penalty: $\{\ell_1, \ell_2\}$, C: $\{0.01 \dots 10\}$)<br>Random Forest (n_estimators: 100, max_depth: 4) |
| Realizations | 100 trials of independent 50/50 train/test splits |

### Intersectional de-biasing improves fairness for small and large groups

It is challenging to build models that both perform well on marginalized groups and minimize overfitting. This is particularly concerning when evaluating intersectional fairness approaches, as with each additional attribute to consider, the number of groups grows factorially while group size decreases. Therefore, we evaluate how the benefits of intersectional de-biasing approaches are distributed across the groups of varying prevalence. In the MIMIC-IV ED, intersectional de-biasing approaches minimize both the group-specific ECE (Fig. 4, top left) and the FNRs for the lowest prevalence group (AIAN, M, prevalence=0.11%) and highest prevalence groups (White, F, prevalence=31.36%), in contrast to no de-biasing, single-attribute de-biasing, and multi-attribute, marginal de-biasing. For intermediate prevalence groups (0.16% to 28.39%), intersectional de-biasing either outperformed or equalled all other de-biasing conditions in the MIMIC-IV data. Similar performance was noted in the pediatric setting across both fairness optimization targets (Fig. 4, bottom left and right).

**Figure 4:**
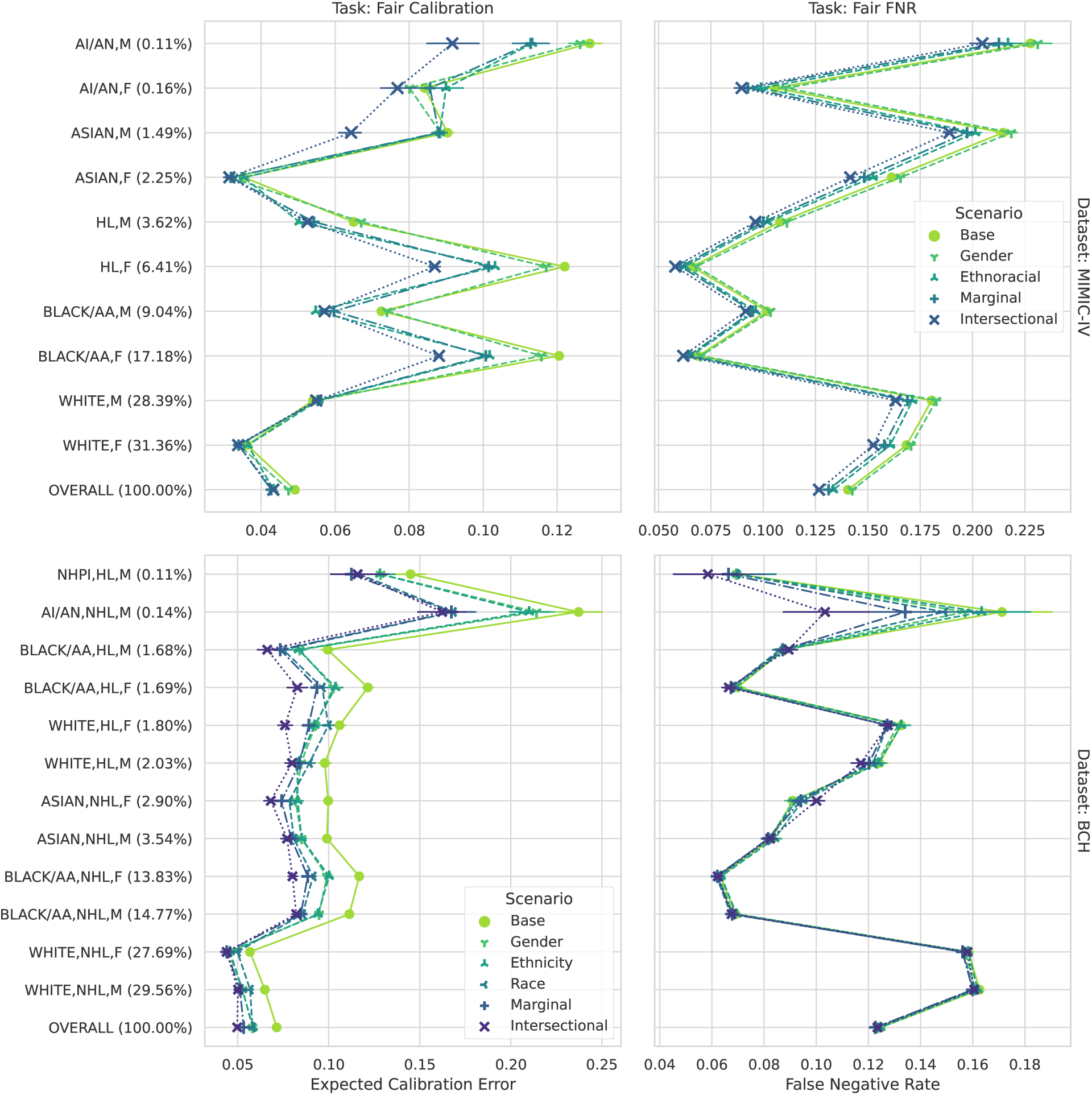
Model performance on each intersectional position (y-axis) according to dataset (top: MIMIC-IV, bottom: BCH), fairness consideration (left: expected calibration error, right: false negative rate), demarcated by scenario. Points indicate bootstrap-estimated median performance over trials and bars indicate the 95% confidence interval. AI/AN: American Indian / Alaskan Native; AA: African American; NHPI: Native Hawaian Pacific Islander; (N)HL: (Not) Hispanic/Latino; F: Female; M: Male.

## DISCUSSION

### Exclusions and Limitations

To date, most model bias is identified post-deployment^40^, with few clinical prediction models incorporating fairness notions in the development process. This study is among the first to implement an intersectional de-biasing approach for clinical prediction models and demonstrate that 1) it can significantly improve the performance of a model on subgroups versus the more common, marginal approaches; and 2) it can reduce unfairness with minor changes in overall performance. In MIMIC-IV, intersectionally de-biased ML models exhibit a 27% reduction in subgroup ECE or 11% reduction in subgroup FNR with no change in AUROC or AUPRC; in BCH, models exhibit a 27% reduction in subgroup ECE with no reduction in AUROC or AUPRC, and a 6.4% reduction in subgroup FNR for no reduction in AUROC and a 3% reduction in mean AUPRC (concentrated at low sensitivity model thresholds).

A challenge of intersectional approaches using demographic traits is that as more protected attributes are added, group sizes shrink. We limited our analysis to three attributes: race, ethnicity, and sex, and only considered intersectional groups representing at least 0.1% of the population. While multicalibration handles small group sizes with a threshold, other fairness methods use a prior probability for group outcomes. We tested both approaches in FOMO and found no significant effect on results. Future studies could explore additional attributes and larger datasets to examine the limits of fairness gains for smaller intersectional groups.

Our results are limited to one clinically relevant prediction problem, but it is a type of resource allocation problem that is widely found in clinical settings. Further work should examine the extent to which our observations generalize to other settings of interest, which may additionally have their own appropriate measures of fairness.

We do not attempt to answer whether subgroup calibration or subgroup FNRs are a more important fairness consideration for this task; instead, we attempt to measure the importance of intersectional de-biasing of multiple scenarios. Calibration is important for interpreting risk scores and doing risk stratification. FNRs are important for interpreting the risk of missed interventions (in this case, hospital admissions). It is well known that FNRs, FPRs, and calibration cannot be simultaneously equal when subgroups exhibit different prevalence of the outcome^33^. Future studies could consider two-way optimizations of these fairness metrics which are not covered here. Similarly, future prospective studies depend on extended engagement with community collaborators to define which metrics are more important in clinical decision support.

## Data Availability

MIMIC-IV-ED is available from physionet.org/mimic-iv-ed. The full preprocessing code for the MIMIC-IV admissions dataset is available from the repository github.com/cavalab/mimic-iv-admissions. The BCH pediatric dataset is not publicly available under the terms of the BCH Institutional Review Board. Interested readers may contact the corresponding author for additional details.

## Code Availability

The code for reproducing the experiments is available from github.com/cavalab/marginal-intersectional.

## Author Contributions

EL and WGL conceived the study and designed the experiment. EL wrote the initial manuscript and contributed to code and experimental evaluation. SS and WGL wrote methods and experimental code and ran the experiments. WGL created the tables and figures and contributed to writing the manuscript. YBC developed and curated the BCH dataset. YBC, AF and BYR provided feedback and guidance on the study design, clinical use case, and manuscript.

## Additional Information

Supplementary Information is available for this paper. Correspondence and requests for materials should be addressed to.

## Acknowledgments

This work was partially supported by National Institutes of Health grant no. R01LM014300 from the National Library of Medicine.

## SUPPLEMENT

### 1 Additional Cohort Details

**Table S1:** Fraction of emergency admissions (%) by intersectional position for patients in the MIMIC-IV (top) and BCH (bottom) cohorts. AI/AN: American Indian / Alaskan Native; AA: African American; NHPI: Native Hawaian Pacific Islander; (N)HL: (Not) Hispanic/Latino; F: female; M: male. Subgroups with fewer than five samples are omitted.

| MIMIC-IV ED |  |  |  |  |
| --- | --- | --- | --- | --- |
| Gender |  | Female | Male | Overall |
| Ethnoracial Group |  |  |  |  |
| AI/AN |  | 70 / 257 (27) | 82 / 170 (48) | 152 / 427 (36) |
| ASIAN |  | 1,043 / 3,595 (29) | 1,032 / 2,384 (43) | 2,075 / 5,979 (35) |
| BLACK/AA |  | 3,124 / 27,486 (11) | 2,603 / 14,458 (18) | 5,727 / 41,944 (14) |
| HL |  | 1,063 / 10,262 (10) | 1,168 / 5,795 (20) | 2,231 / 16,057 (14) |
| WHITE |  | 18,147 / 50,174 (36) | 18,951 / 45,435 (42) | 37,098 / 95,609 (39) |
| Overall |  | 23,447 / 91,774 (26) | 23,836 / 68,242 (35) | 47,283 / 160,016 (30) |
| BCH ED |  |  |  |  |
| Race | Gender Ethnicity | Female | Male | Overall |
| AI/AN | HL | - | - | - |
|  | NHL | 1 / 5 (20) | 10 / 19 (53) | 11 / 24 (46) |
|  | Overall | 1 / 7 (14) | 10 / 21 (48) | 11 / 28 (39) |
| ASIAN | HL | - | - | - |
|  | NHL | 61 / 398 (15) | 74 / 484 (15) | 135 / 882 (15) |
|  | Overall | 61 / 401 (15) | 74 / 487 (15) | 135 / 888 (15) |
| BLACK/AA | HL | 22 / 232 (9) | 25 / 231 (11) | 47 / 463 (10) |
|  | NHL | 188 / 1,892 (10) | 212 / 2,028 (10) | 400 / 3,920 (10) |
|  | Overall | 210 / 2,124 (10) | 237 / 2,259 (10) | 447 / 4,383 (10) |
| NHPI | HL | 2 / 9 (22) | 1 / 15 (7) | 3 / 24 (12) |
|  | NHL | 2 / 7 (29) | 1 / 4 (25) | 3 / 11 (27) |
|  | Overall | 4 / 16 (25) | 2 / 19 (11) | 6 / 35 (17) |
| Other | HL | 261 / 2,812 (9) | 308 / 2,963 (10) | 569 / 5,775 (10) |
|  | NHL | 182 / 1,232 (15) | 247 / 1,500 (16) | 429 / 2,732 (16) |
|  | Overall | 443 / 4,044 (11) | 555 / 4,463 (12) | 998 / 8,507 (12) |
| WHITE | HL | 53 / 247 (21) | 45 / 280 (16) | 98 / 527 (19) |
|  | NHL | 905 / 3,800 (24) | 1,017 / 4,054 (25) | 1,922 / 7,854 (24) |
|  | Overall | 958 / 4,047 (24) | 1,062 / 4,334 (25) | 2,020 / 8,381 (24) |
| Overall | HL | 338 / 3,305 (10) | 379 / 3,494 (11) | 717 / 6,799 (11) |
|  | NHL | 1,339 / 7,334 (18) | 1,561 / 8,089 (19) | 2,900 / 15,423 (19) |
|  | Overall | 1,677 / 10,639 (16) | 1,940 / 11,583 (17) | 3,617 / 22,222 (16) |

Table S1 shows a detailed breakdown of patient admission characteristics over combinations of race, ethnicity and gender.

### 2 Additional Experiment Details

#### Data preprocessing and cleaning

For numeric data in the MIMIC-IV-ED triage table (Table 3), we encoded outliers as NaNs according to the following (min,max) ranges: temperature (95-105 F); heart rate (30-300 beats per minute); respiratory rate (2-200 breaths per minute), oxygen saturation (50-100%); systolic blood pressure (30-400 mmHg), diastolic blood pressure (30-300 mmHg); pain scores (0-20); acuity score (1-5).

For both cohorts, chief complaint consists of brief strings of free text. For these data, we first applied simple harmonization and cleaning heuristics and then one-hot- encoded the result, filtering out tokens ocurring less than 1% of the time. In our preliminary analysis we evaluated the use of pre-trained word embeddings for chief complaint but did not find that they improved performance versus one-hot-encoding.

#### Algorithm Implementation

We use a Python implementation of Multicalibration Boosting available from github.com/cavalab/pmcboost and derived from La Cava, Lett, and Wan [41]. Fairness-Oriented Multiobjective Optimization (FOMO) is available from cavalab.org/fomo. FOMO serves as a generic interface between the multi-objective optimization algorithms from [pymoo](https://pymoo.org/) and ML methods that follow the scikit-learn API while accepting sample weights as an argument during training (i.e. in calls to fit()). Our experimental study focuses on utilizing the popular NSGA2^42^ algorithm in conjunction with two widely used ML methods that support weighted classification: random forests (implemented in [XGBoost](https://xbgoost.org/)) and penalized linear regression (implemented in scikit-learn^43^). The code to run the experiments is available from the repository github.com/cavalab/marginal-intersectional.

#### Training

We ran 100 trials of each combination of dataset (MIMIC-IV, BCH), fairness task (fair calibration, fair false negative rates), group construction scenario (Base, Race, Gender, Ethnicity, Marginal, Intersectional), and base model (penalized logistic regression, random forests), as shown in Table 4. Each trial utilized a unique random seed that resulted in a random shuffle of the data which was split into 50% train/ 50% test sets. Splits were stratified by outcome (admission), gender, and race to maintain appropriate representation in each. For the runs using FOMO and MIMIC-IV data, the training set was further reduced to 10% (approximately 16k patients) to reduce computation time.

### 3 Additional Experiments

In this section we report additional experiments meant to characterize the sensitivity of the studied fairness algorithms to hyperparameters and design variables. For both multicalibration boosting and FOMO, we analyze how the choice of base ML model, group prevalence, and dataset affect the results. In the case of multicalibration boosting, we studied the choice of *α*, a termination criteria that defines the group-specific calibration error threshold, and *γ*, a parameter that controls the minimum prevalence of a group to be considered for updating. In the case of FOMO, we looked at the effect of using a weighted subgroup FNR metric that accounts for prior probability of the groups, and the effect of a fairness meta-model complexity.

#### 3.1 Multicalibration Boosting

##### Sensitivity Analysis

In Fig. S1, we visualize the expected calibration error of LR and RF models on MIMIC-IV as a function of base model, *α*, *γ*, and modeling scenario. At higher levels of *γ*, low-prevalence groups are excluded from fairness updating; hence, performance differences between scenarios tend to shrink. Relatedly, higher values of *α* loosen the threshold needed for multicalibration to perform an update, and so model performance tends to become similar between groups. Conversely, for very small values of *α* and *γ*, small groups have a larger impact on fairness optimization, meaning intersectional modeling matters more for achieving low ECE among intersectional groups. Overfitting can occur when *α* is too stringent, leading to degradation of performance on intersectional groups on test set: see top middle and right panel of Fig. S1, RF models.

**Figure S1:**
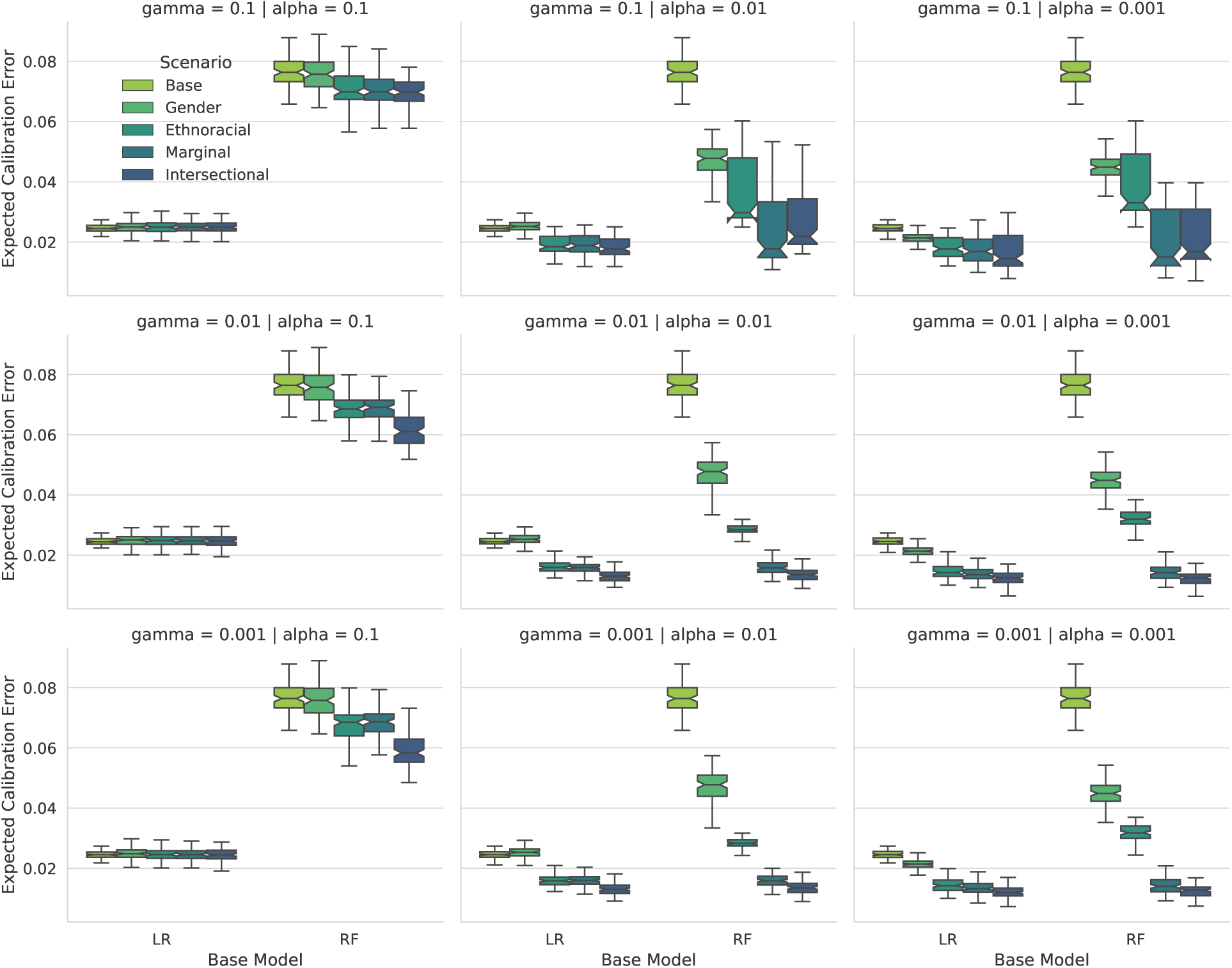
Intersectional group-wise Expected Calibration Error on MIMIC-IV as a function of *γ* (row), *α* (column), base ML model (x-axis), and optimization scenario (color). At high levels of *α*, the models remain unchanged, whereas at very low values of *α* and *γ*, performance on intersectional groups can suffer due to small sample sizes.

Fig. S2 sheds light on the interaction between group prevalence, *α* and *γ* under multicalibration boosting. Here we explicitly look at training and test set performance of the intersectional de-biasing approach relative to the baseline approach, illustrating how the constraints on calibration error (*α*) and minimimum group probability (*γ*) interplay with group prevalence (x-axis). In general, we observe that groups that are less prevalent in the data tend to have higher expected calibration error (ECE). Therefore, when *α* and *γ* are set high relative to model performance on adequately sized groups (e.g., *α* = *γ* = 0.1, top left panel), no de-biasing occurs. Conversely, if *γ* and *α* is set very low, de-biasing occurs over all groups in the training data but this does not fully generalize to test data (bottom right panel).

**Figure S2:**
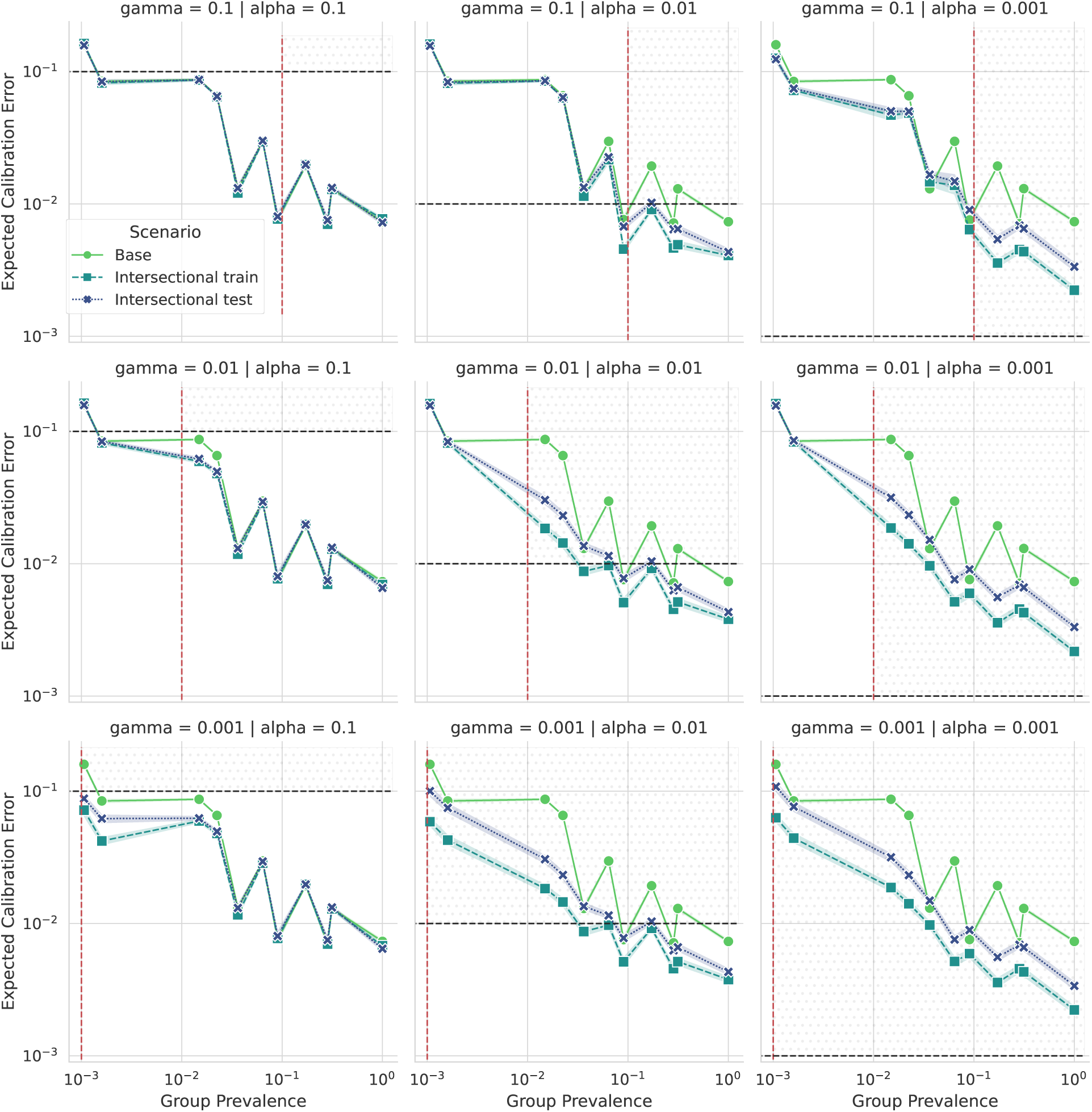
Expected calibration error (ECE) as a function of group prevalence for LR models trained on MIMIC-IV, under different combinations of *α* and *γ*. The shaded area indicates the region of model performance that is subjected to optimization by either having an ECE higher than the threshold, *α*, or a group prevalence higher than the cutoff, *γ*.

**Figure S3:**
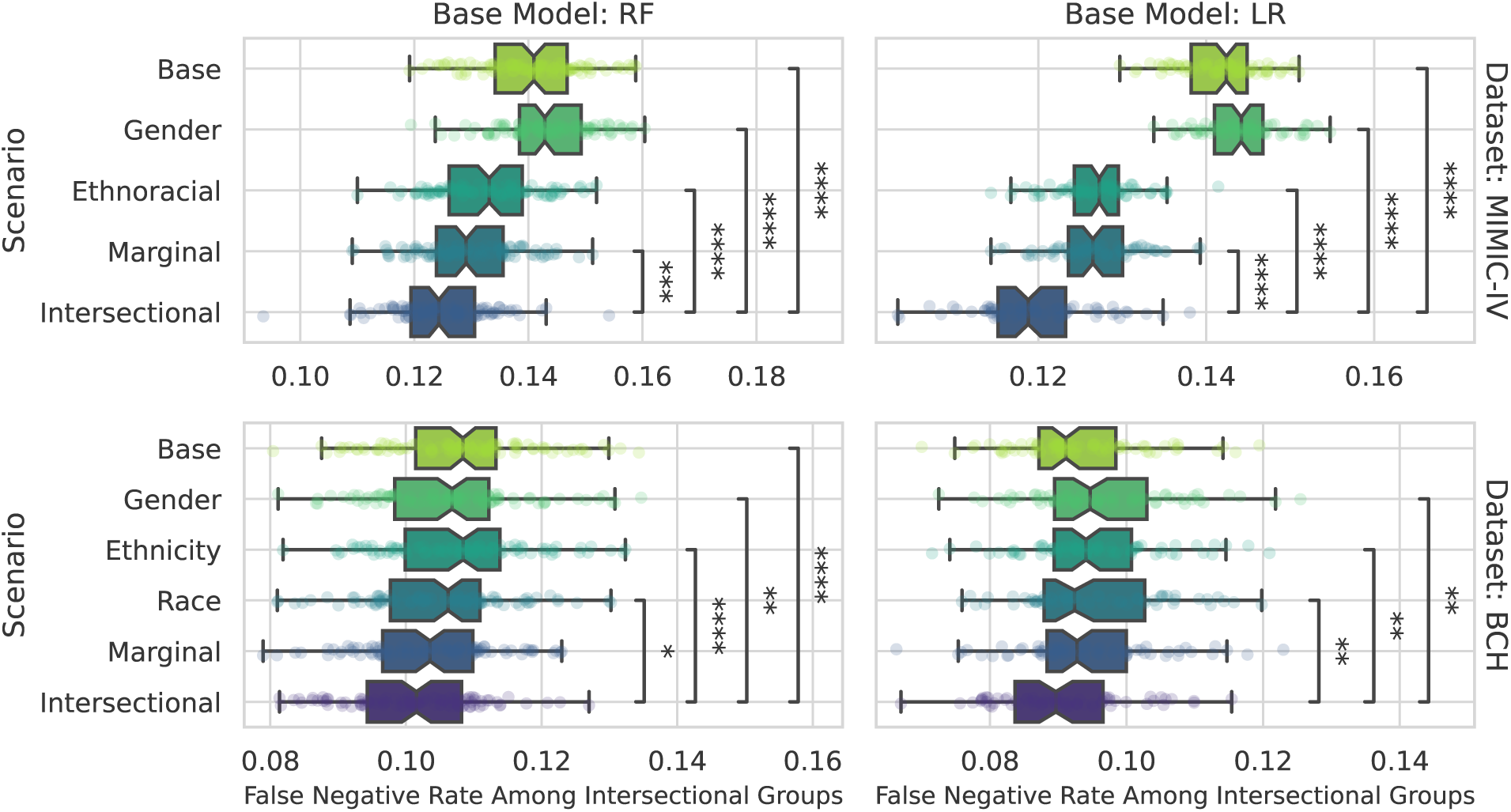
False negative rates (FNR) among intersectional groups under different base models (left: random forests (RF), right: penalized logistic regression (LR)) and FOMO de-biasing scenarios (y-axis) for MIMIC-IV (top) and BCH (bottom). Statistical tests are two-sided Mann-Whitney-Wilcoxon tests with Holm-Bonferroni correction ( *: 1e-2 < p <= 5e-2; **: 1e-3 < p <= 1e-2; ***: 1e-4 < p <= 1e-3; ****: p <= 1.0e-4).

#### 3.2 Fairness-Oriented Multiobjective Optimization

##### Sensitivity Analysis

We varied several parameters during our experimentation with FOMO: 1) The choice of ML model (penalized logistic regression or random forests); 2) whether the definition of subgroup fairness incorporates the prior probability of the group as in other work^13^; 3) the type of meta-model used to estimate the sample weights used to train the base models. Regarding 1), we saw similar trends in results when working with linear models, as shown in Fig. S3. Regarding 2), we did not observe a difference in performance when incorporating prior probabilities of the groups; our results here do not incorporate these adjustments for group size. Regarding 3), we did not observe a difference in performance with variations of the meta-model. In our results, we use a standard linear formulation to map patient attributes to training sample weights; when using the intersectional fairness implementation, we extend the linear model with interaction terms between the scenario’s protected features. Our observations suggest that whether or not the group probability was factored into the fairness definition, it had minimal discernible impact on the outcomes for both RF and LR models across both datasets.

##### Trade-off Visualization

Fig. S4 shows the set of models generated by FOMO as part of its optimization process, which characterizes the trade-off space (i.e. the Pareto frontier) between fairness and accuracy objectives.

**Figure S4:**
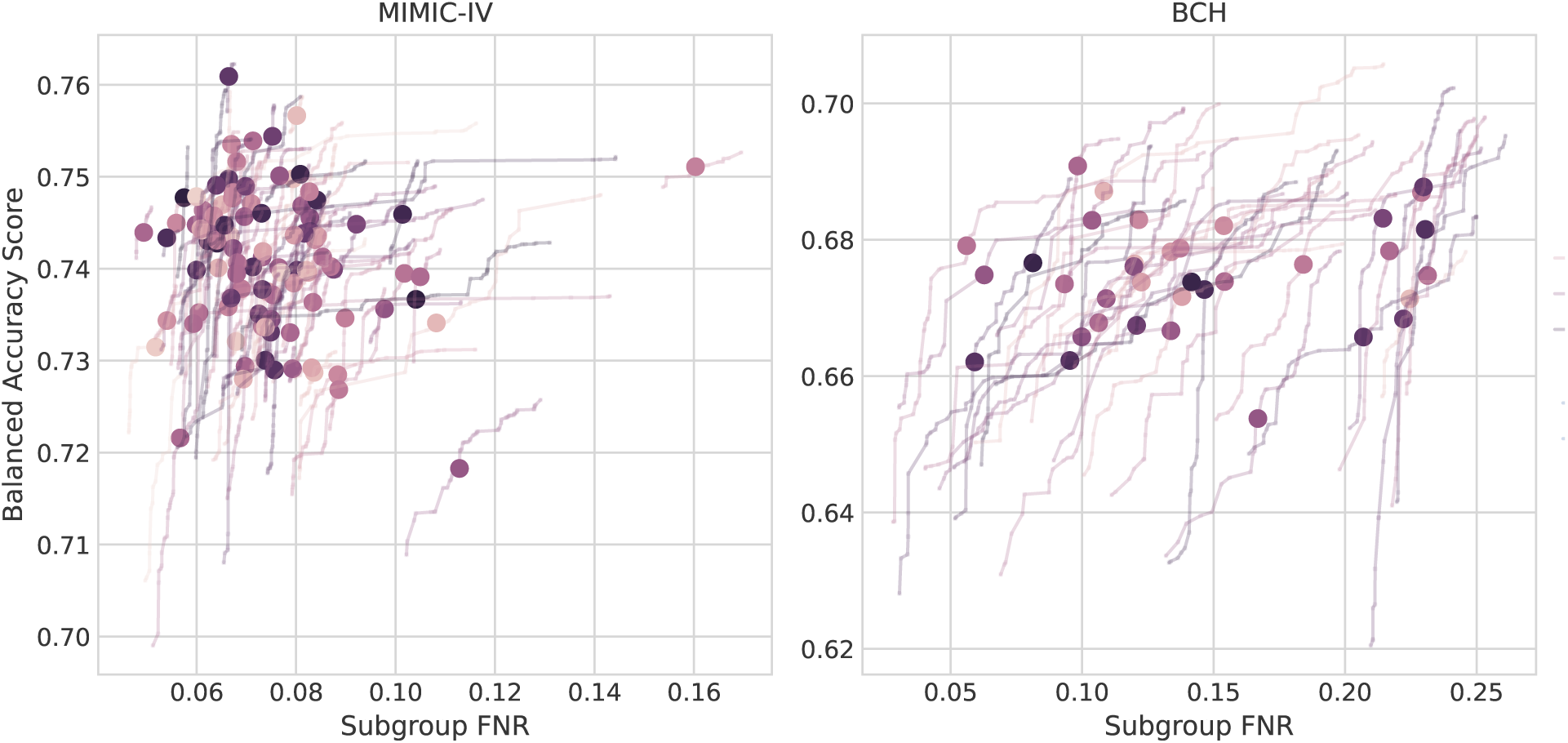
Accuracy-Fairness Tradeoffs and Model Selection. FOMO optimizes a Pareto frontier of solutions simultaneously in order to characterize the trade-off between accuracy and fairness objectives. These final frontiers are shown for MIMIC-IV (left) and BCH (right), with each line representing one realization of the experiment. In order to choose a final model (marked by large circles for each run), a multi-criteria decision making method known as Pseudo-Weights is used^42^. This method chooses the model that maximizes a weighted sum of the objectives. For each candidate model, the weights of each objective depend on the normalized distance to the worst solution for that objective. FNR: false negative rate.

